# Application of COVID-19 pneumonia diffusion data to predict epidemic situation

**DOI:** 10.1101/2020.04.11.20061432

**Authors:** Wu zhenguo

## Abstract

**Objective:** To evaluate novel coronavirus pneumonia cases by establishing the mathematical model of the number of confirmed cases daily, and to assess the current situation and development of the epidemic situation, so as to provide a digital basis for decision-making.

**Methods:** The number of newly confirmed covid-19 cases per day was taken as the research object, and the seven-day average value (**M**) and the sequential value (**R**) of M were calculated to study the occurrence and development of covid-19 epidemic through the analysis of charts and data.

**Results:** **M** reflected the current situation of epidemic development; **R** reflected the current level of infection and the trend of epidemic development.

**Conclusion:** The current data can be used to evaluate the number of people who have been infected, and when **R** < 1, the peak of epidemic can be predicted.

**Preface:** In December 2019, a number of cases of pneumonia with unknown causes were found in some hospitals in Wuhan, Hubei province, China. On 11 March 2020, the director-general of the world health organization (WHO), Tedros Adhanom Ghebreyesus, announced that based on the assessment, WHO believes that the current outbreak of COVID-19 can be called a global pandemic. By early April 2020, there were more than one million confirmed cases worldwide.

COVID-19 has developed from sporadic cases to pandemic in a short period of 3 months. The analysis and research of its infectious data will help to prevent and control the next stage of epidemic prevention and other infectious diseases in the future.

In this paper, COVID-19 rounded average of seven days (**M**), and M’s ring ratio (**R**) are used to predict the current potential patients’ data, and the relative state of epidemic prevention and control is judged through the graphic features and characteristic data, so as to provide evidence for the prevention and control decisions.

## 1. Object and method

### 1.1 research object

The number of confirmed COVID-19 cases which increased daily.

### 1.2 investigation method

#### 1.2.1 main analysis objects

The report of epidemic data is affected by many factors, such as insufficient detection kits and insufficient personnel matching, and the new data in a single day fluctuates greatly. We selected the multi-day moving average (**M**_n_) of the number of newly confirmed COVID-19 cases per day as the main analysis object.

#### 1.2.2 daily moving average M_n_ of newly confirmed cases (n is a set constant)

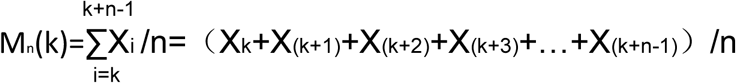

#### Definition

X_1_ is the new increment of the first day, X_i_ is the new increment of the first day since data record.

In the operation, M_n_(k) will be rounded and start on the nth day(k≥n.)

#### 1.2.3 diffusion ratio (the ratio of Mn in n days) R_n_ (n is the set constant)

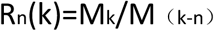

In the operation, R_n_(k) will be rounded and start on the 2n day (K ⩾ 2n).

##### Definition

Diffusivity Rn describes the average relative relationship between newly added patients in recent n days and newly added patients in the previous group in n days, which reflects the level of infection to some extent.

In extreme cases, assuming that all the patients with the most recent n day onset of an infectious disease were infected by the last n day patient, the value of R_n_ basically corresponds to the basic infectious number (R_0_) in epidemiology.

### 1.2.4 value range of R

Due to the influence of detection, statistics and other factors, the **M**_n_ value may jump from a lower level to a higher level, and the calculation value of the diffusion ratio **R** may be very large. At this time, it does not reflect the current level of infection. We limit the maximum value of R to reduce the impact on the overall operation.

We define a maximum **R** of 5 (refer to the higher level of infectious diseases R_0_) to avoid excessive fluctuations in the data:

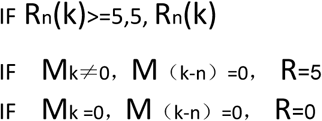

In epidemiology, the basic infection number is the average number of people who will be infected with an infectious disease and spread the disease to others without external intervention or immunity.

#### 1.2.5 estimated number of potential patients (n is a set constant)

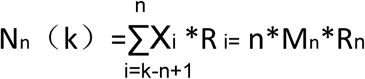

That is, the estimated number of potential patients on the i-th day is equal to the cumulative value of the new data (including the i-th day) on the previous n-th day multiplied by the diffusion rate on that day. In other words, assuming the development of the epidemic maintains the current infection rate, the number of potential cumulative patients in the next cycle (n days).

#### 1.2.6 Estimates of the total number of infections

We estimate the total number of infections through the current cumulative number of infections plus the number of potential patients, so that we can roughly assess the scale and trend of the current epidemic.

Estimates of the total number of infections

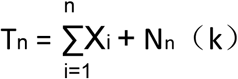

## 2. Data analysis and Application

### 2.1 data assumption of outbreak

In addition to the cluster infection caused by specific cases (Diamond Princess), more than 100 cases were added in a single day, indicating that there have been multiple community infections in a region, and the outbreak has occurred on a certain scale.

### 2.2 n-day moving average of newly confirmed cases **M**_n_

The moving average helps to eliminate data distortion caused by statistics or other human factors. The relationship between the moving average value Mn formed by different short, medium and long periods (n) and the formation of original data can be used to judge the stage of epidemic situation.

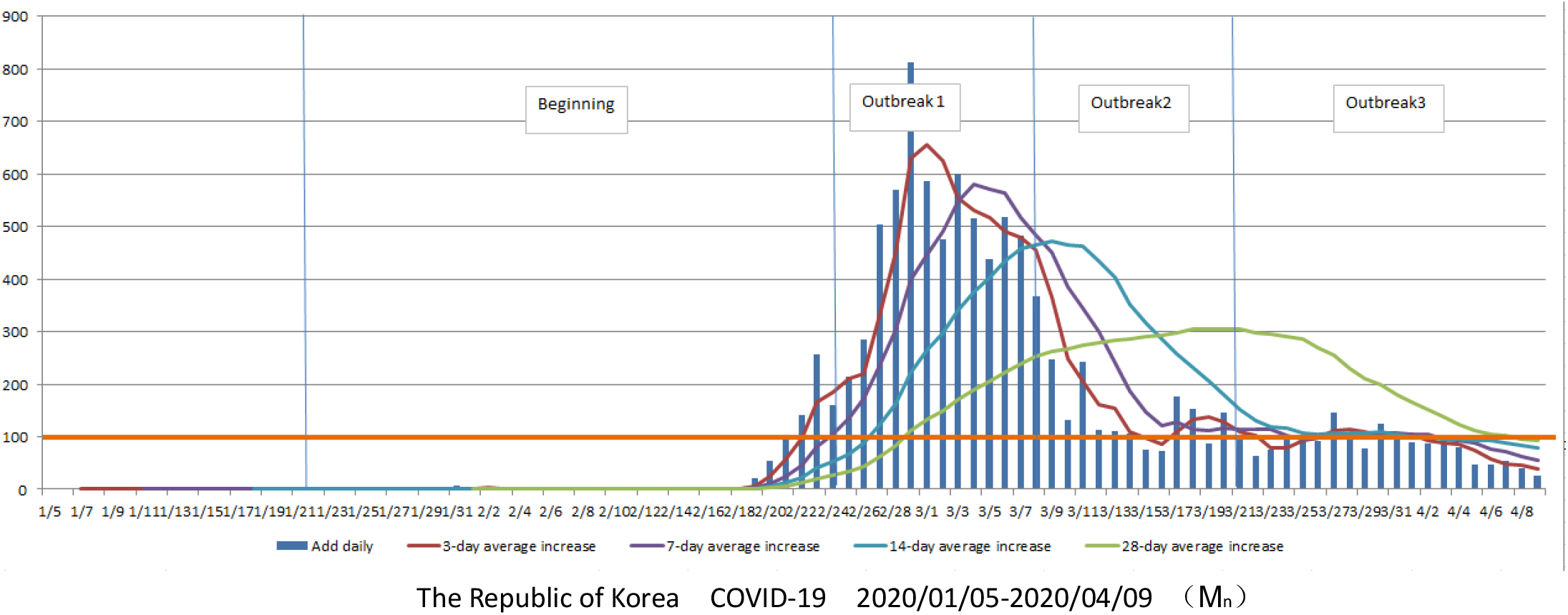

| Features | Add daily | 3-day<br>average<br>increase<br>$M_3$ | 7-day<br>average<br>increase<br>$M_7$ | 14-day<br>average<br>increase<br>$M_{14}$ | 28-day<br>average<br>increase<br>$M_{28}$ |
| --- | --- | --- | --- | --- | --- |
| Beginning | $<100$ | | | | |
| Outbreak 1 | $\geq 100$ | $\geq 7$-day<br>average<br>increase | $\geq 14$-day<br>average<br>increase | $\geq 28$-day<br>average<br>increase | |
| Outbreak 2 | $\geq 100$ | $< 7$-day<br>average<br>increase | | | |
| Outbreak 3 | | | | $\leq 28$-day<br>average<br>increase | |

Features of COVID-19 ( $M_n$ )

### 2.3 diffusion ratio (ratio of **M**_n_ in n days) **R**_n_

In the early stage of the outbreak in South Korea (from January 5, 2020 to February 20, 2020), R_3_, R_5_ and R_7_ fluctuated irregularly between 0-2. After February 20, 2020 (add Daily > 100), R_3_, R_5_ and R_7_ exceeded the limits respectively (according to the definition above, value 5), showing an outbreak trend, then quickly fell back to 1, and continued to April 9, 2020, the epidemic situation was in a medium development level.

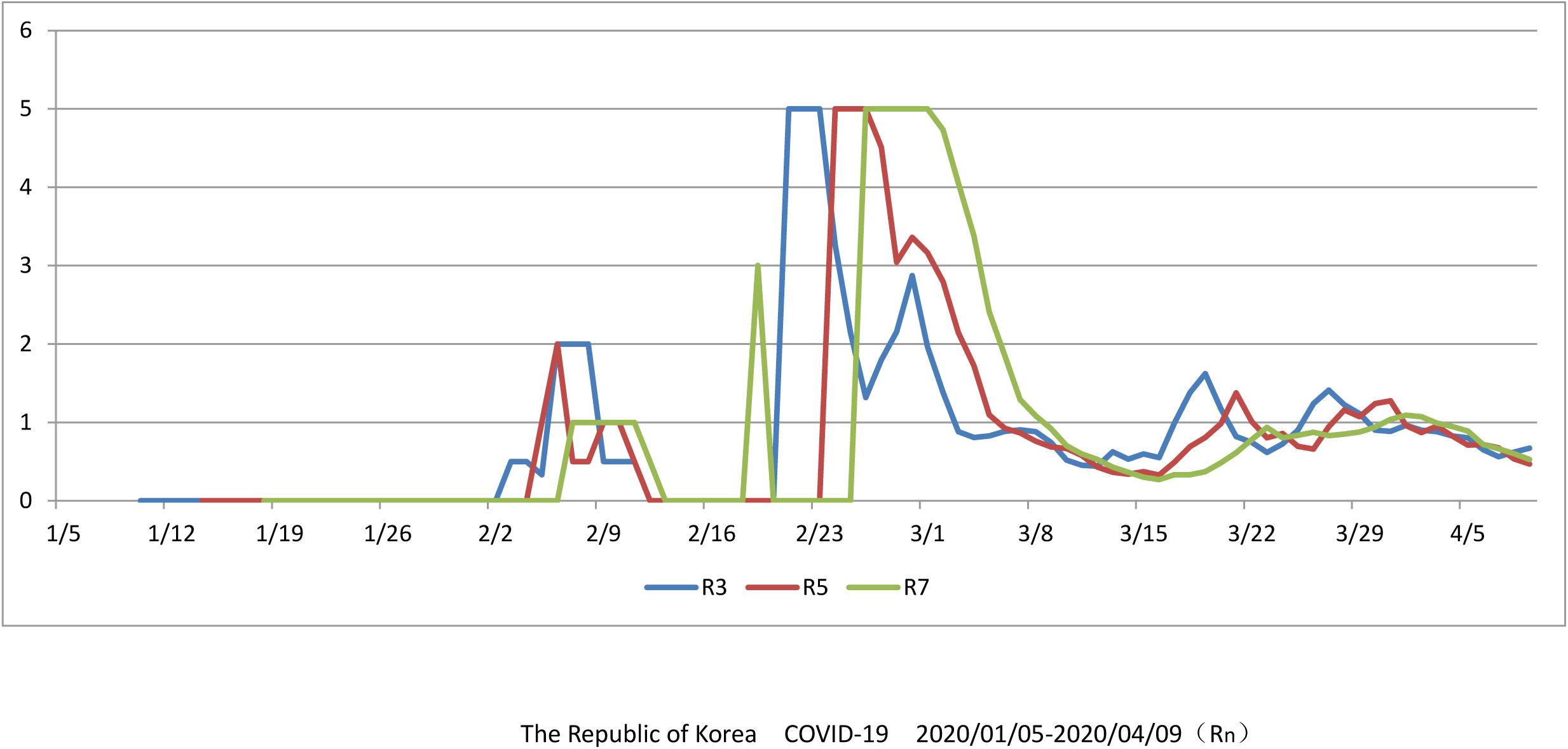

From January 5, 2020 to April 4, 2020, R_3_, R_5_, and R_7_ in Singapore showed wide fluctuations and jagged graphs. The number of new cases in a single day never exceeded 100. The outbreak was in the early stage. On April 5, 2020, Singapore entered the outbreak phase with more than 100 new cases in a single day.

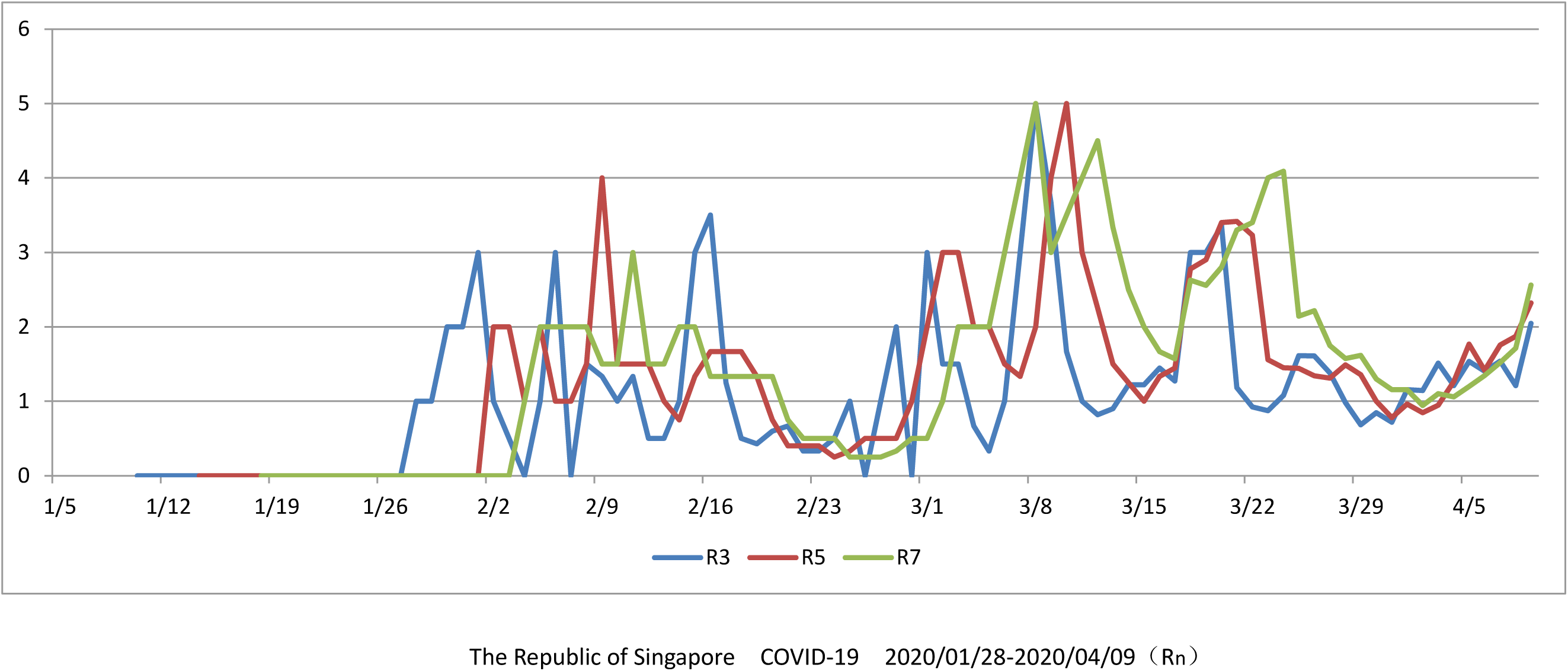

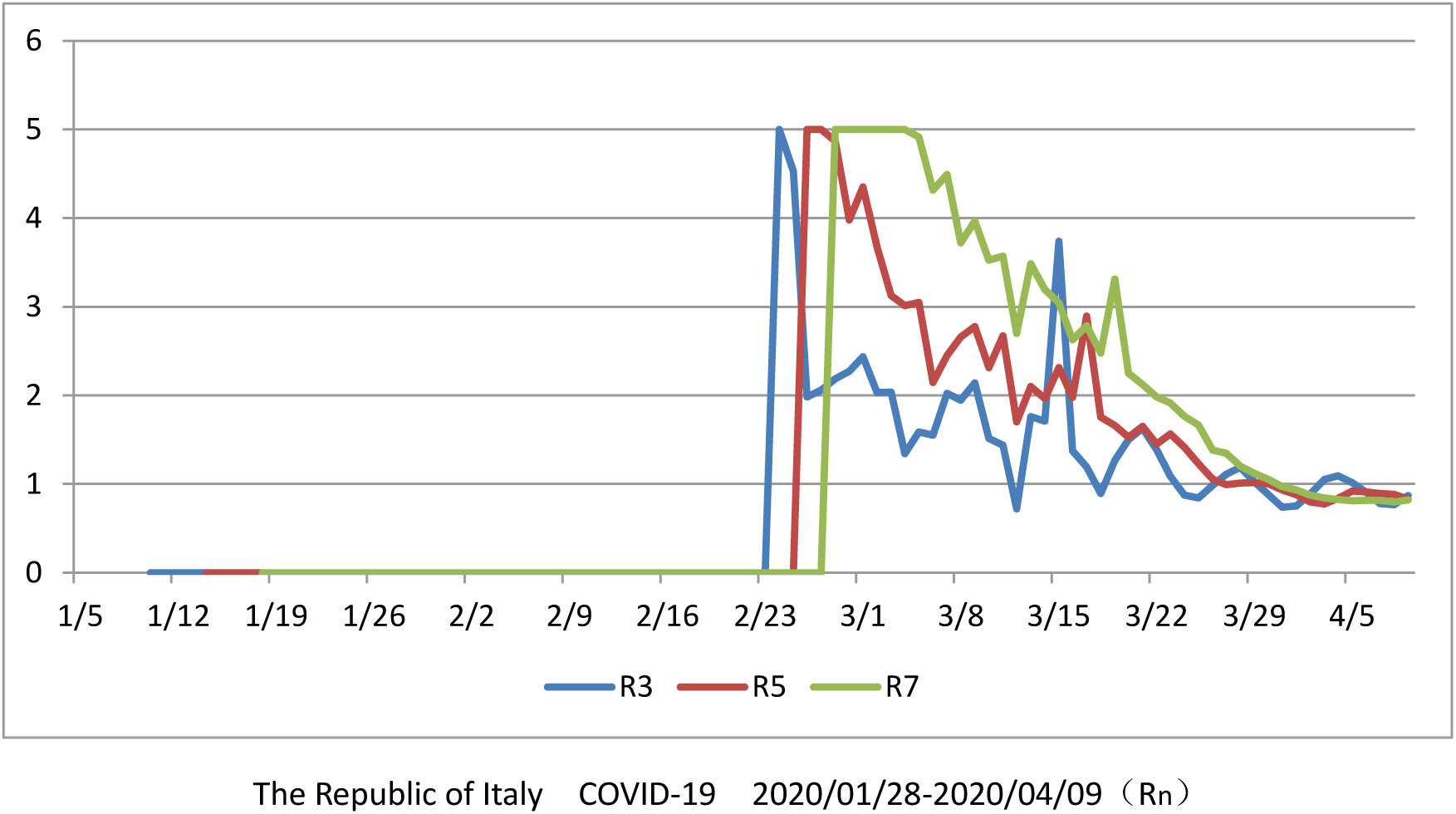

From February 21, 2020 to April 9, 2020, Italy R_3_, R_5_ and R_7_ exceeded the limits respectively (according to the above definition, value 5), and then gradually fell back slowly, being more than 1 for a long time, in an explosive state. After March 31, 2020, R_3_, R_5_ and R_7_ fell back to the fluctuation near 1, and the epidemic is now in a medium development level.

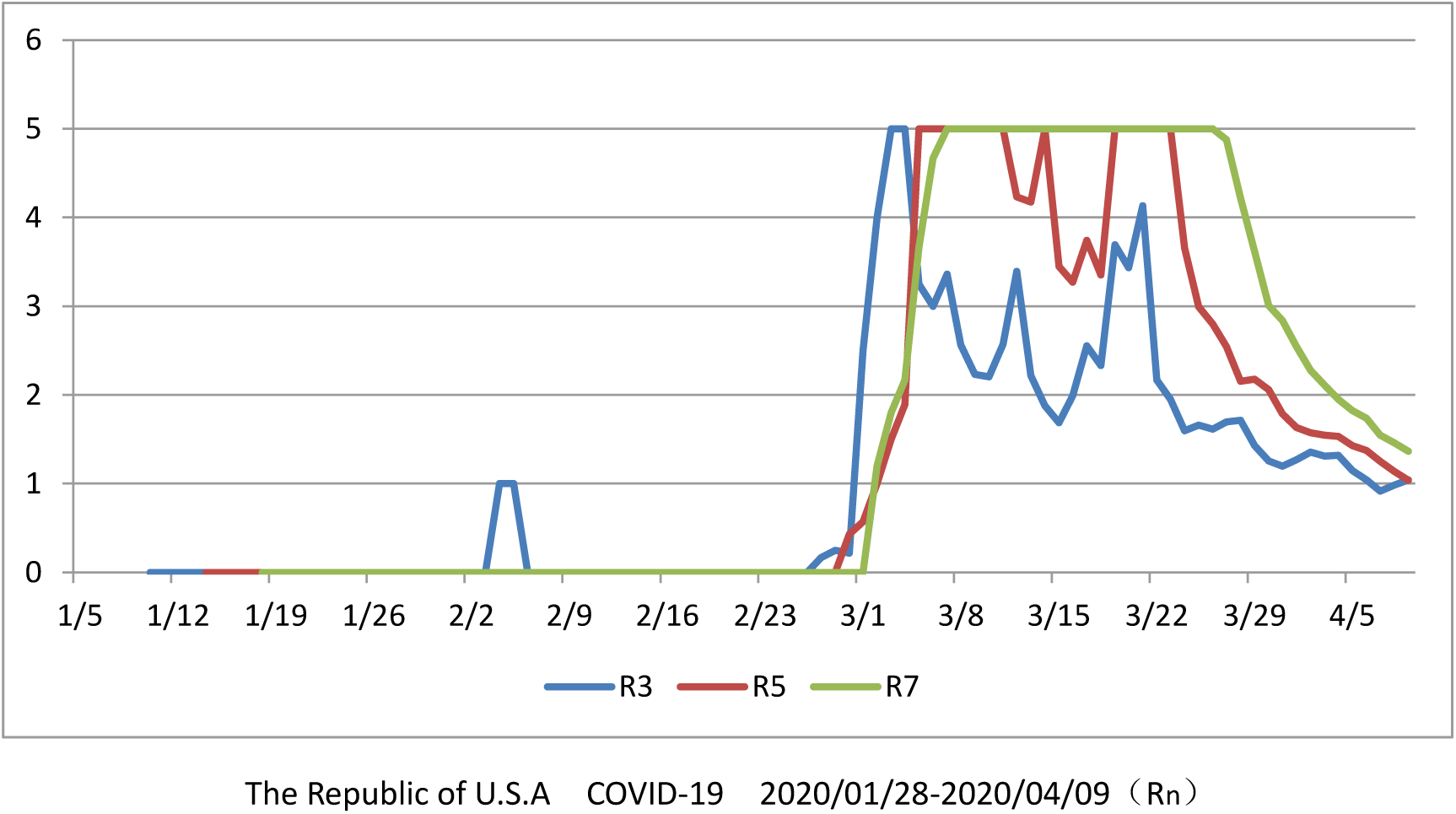

From January 5, 2020 to March 5, 2020, the data of the United States is sporadic and irregular. After March 5, 2020, R_3_, R_5_ and R_7_ are out of the limit range (value 5 according to the previous definition), and remain for a long time (R7 = 5 for 20 days). Compared with other countries, this stage is largely due to the lack of early detection and data concentration release.

From March 27, 2020 to April 9, 2020, R_3_, R_5_ and R_7_ dropped respectively, but remained above 1, and the epidemic was still in the outbreak stage.

### 2.3 n value of diffusion ratio R_n_

When the outbreak is in the early stage, the R_n_ value often continues to reach the upper limit (according to the agreement R_n_ = 5), so the transmission status of the epidemic can not be clearly assessed. Take the Iranian data from January 5, 2020 to April 3, 2020 as an example, R_14_ and R_28_ are equal to the upper limit value for more than 10 days since the calculable date due to the long cycle, which is of little significance for the study and judgment.

In consideration of eliminating the influence of weekends on data statistics, this paper chooses R_7_ as the main reference data for subsequent analysis.

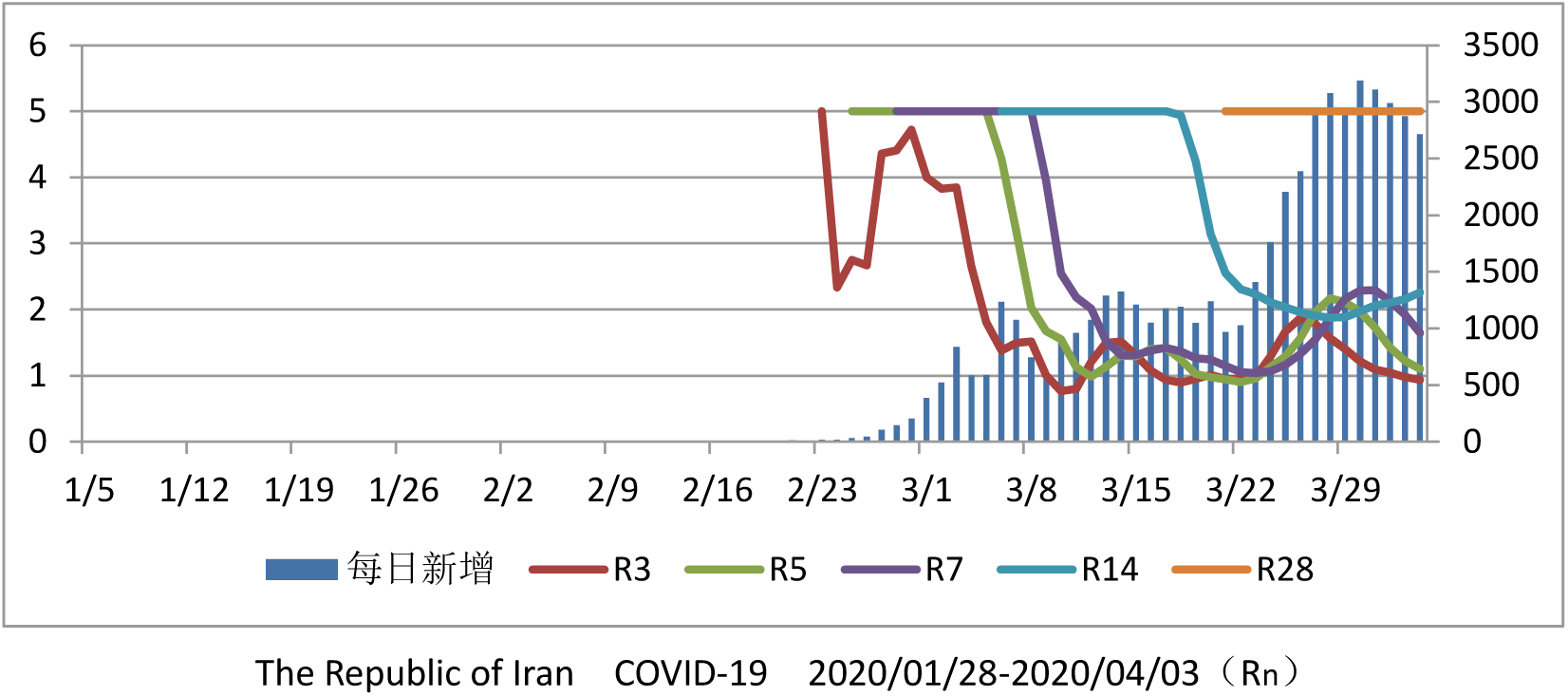

### 2.4 R_7_ distribution in recent week in some countries

R_7_ is the ratio of the number of new infections in a week, which can clearly reflect the degree of infection.

When R_7 (k)_ ⩾ 1, the number of new infections per week will continue to increase, and the epidemic situation will continue;

When R_7 (k)_ is less than 1, the number of new weekly infections continues to fall, and the epidemic situation is gradually controllable.

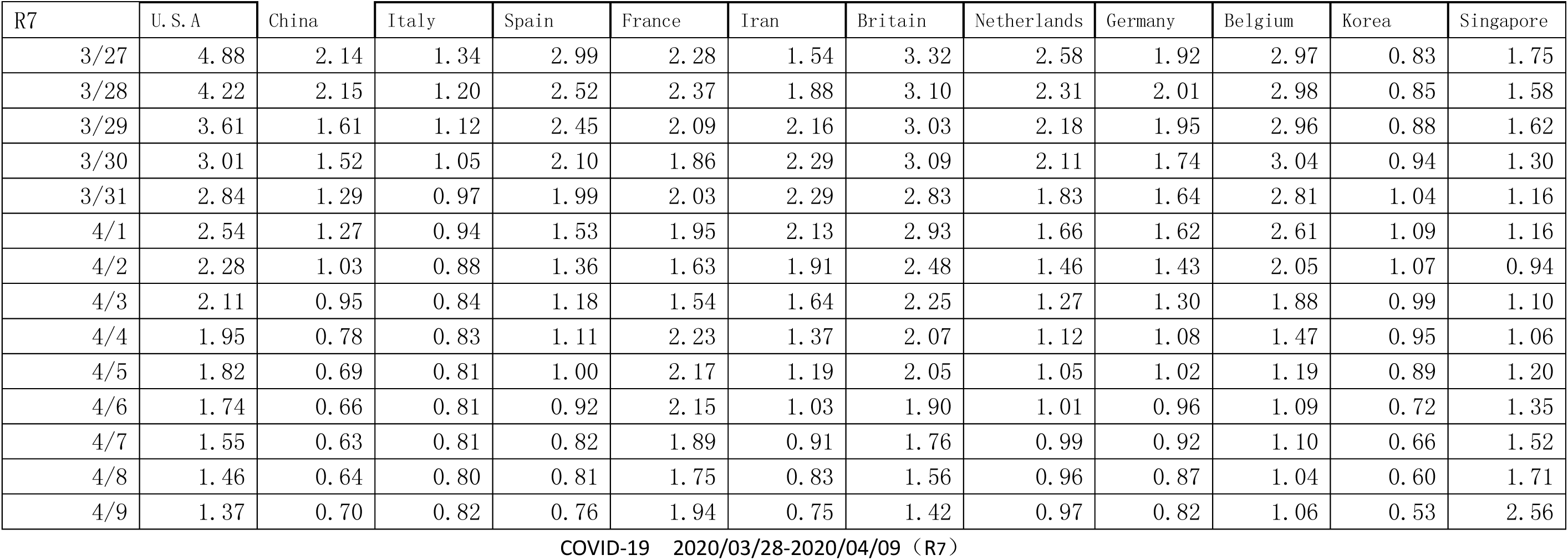

| R7 | U. S. A | China | Italy | Spain | France | Iran | Britain | Netherlands | Germany | Belgium | Korea | Singapore |
| --- | --- | --- | --- | --- | --- | --- | --- | --- | --- | --- | --- | --- |
| 3/27 | 4.88 | 2.14 | 1.34 | 2.99 | 2.28 | 1.54 | 3.32 | 2.58 | 1.92 | 2.97 | 0.83 | 1.75 |
| 3/28 | 4.22 | 2.15 | 1.20 | 2.52 | 2.37 | 1.88 | 3.10 | 2.31 | 2.01 | 2.98 | 0.85 | 1.58 |
| 3/29 | 3.61 | 1.61 | 1.12 | 2.45 | 2.09 | 2.16 | 3.03 | 2.18 | 1.95 | 2.96 | 0.88 | 1.62 |
| 3/30 | 3.01 | 1.52 | 1.05 | 2.10 | 1.86 | 2.29 | 3.09 | 2.11 | 1.74 | 3.04 | 0.94 | 1.30 |
| 3/31 | 2.84 | 1.29 | 0.97 | 1.99 | 2.03 | 2.29 | 2.83 | 1.83 | 1.64 | 2.81 | 1.04 | 1.16 |
| 4/1 | 2.54 | 1.27 | 0.94 | 1.53 | 1.95 | 2.13 | 2.93 | 1.66 | 1.62 | 2.61 | 1.09 | 1.16 |
| 4/2 | 2.28 | 1.03 | 0.88 | 1.36 | 1.63 | 1.91 | 2.48 | 1.46 | 1.43 | 2.05 | 1.07 | 0.94 |
| 4/3 | 2.11 | 0.95 | 0.84 | 1.18 | 1.54 | 1.64 | 2.25 | 1.27 | 1.30 | 1.88 | 0.99 | 1.10 |
| 4/4 | 1.95 | 0.78 | 0.83 | 1.11 | 2.23 | 1.37 | 2.07 | 1.12 | 1.08 | 1.47 | 0.95 | 1.06 |
| 4/5 | 1.82 | 0.69 | 0.81 | 1.00 | 2.17 | 1.19 | 2.05 | 1.05 | 1.02 | 1.19 | 0.89 | 1.20 |
| 4/6 | 1.74 | 0.66 | 0.81 | 0.92 | 2.15 | 1.03 | 1.90 | 1.01 | 0.96 | 1.09 | 0.72 | 1.35 |
| 4/7 | 1.55 | 0.63 | 0.81 | 0.82 | 1.89 | 0.91 | 1.76 | 0.99 | 0.92 | 1.10 | 0.66 | 1.52 |
| 4/8 | 1.46 | 0.64 | 0.80 | 0.81 | 1.75 | 0.83 | 1.56 | 0.96 | 0.87 | 1.04 | 0.60 | 1.71 |
| 4/9 | 1.37 | 0.70 | 0.82 | 0.76 | 1.94 | 0.75 | 1.42 | 0.97 | 0.82 | 1.06 | 0.53 | 2.56 |

COVID-19 2020/03/28-2020/04/09 (R7)

From March 28, 2020 to April 9, 2020, R_7_ in China, Italy, Spain, South Korea and other countries was less than 1, and the epidemic was gradually under control.

The United States, France, the United Kingdom and Belgium are at high levels (R_7_≥ 1), and the epidemic will continue.

### 2.5 estimated dynamic curve of total number of epidemic infections

When the diffusion ratio R > 1, the number of new population will fluctuate and increase, the estimated value of total number of epidemic infections will continue to rise, the epidemic is in the outbreak period (or rising period), and the curve cannot converge.

When the diffusion ratio R < 1, the number of new population will fluctuate and decrease, and the estimated value of total number of epidemic infections will tend to be stable, reflecting that the epidemic situation has been temporarily controlled.

R < 1 is a necessary and sufficient factor for the convergence of the estimated curve of total number of infectious diseases, and it is also an important sign for the epidemic from outbreak to control.

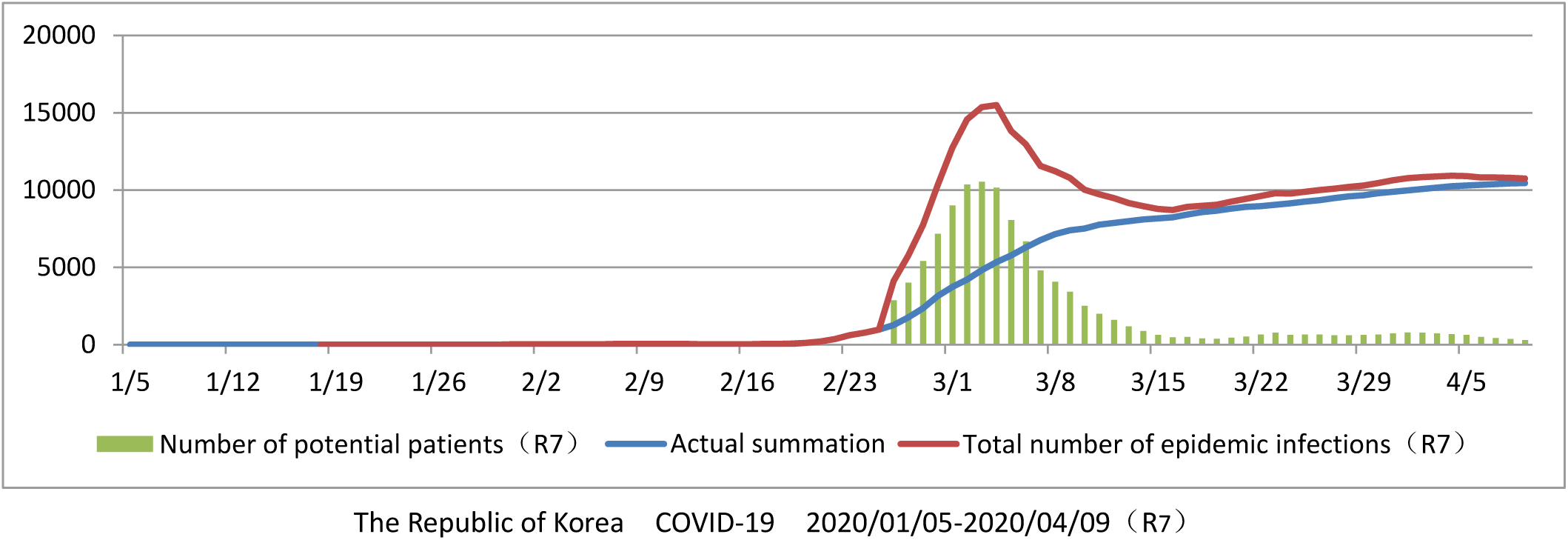

The estimated curve of total number of epidemic infections rose sharply, which was the initial stage of the outbreak.

The estimated value curve of total number of epidemic infections dropped and the epidemic situation was gradually controlled.

The estimated curve of total number of epidemic infections tends to be flatten, and the epidemic situation will be further improved.

The estimated curve of total number of epidemic infections indicates the total number of patients in the future, which can be used as an important basis for medical supplies, equipment and personnel reserves.

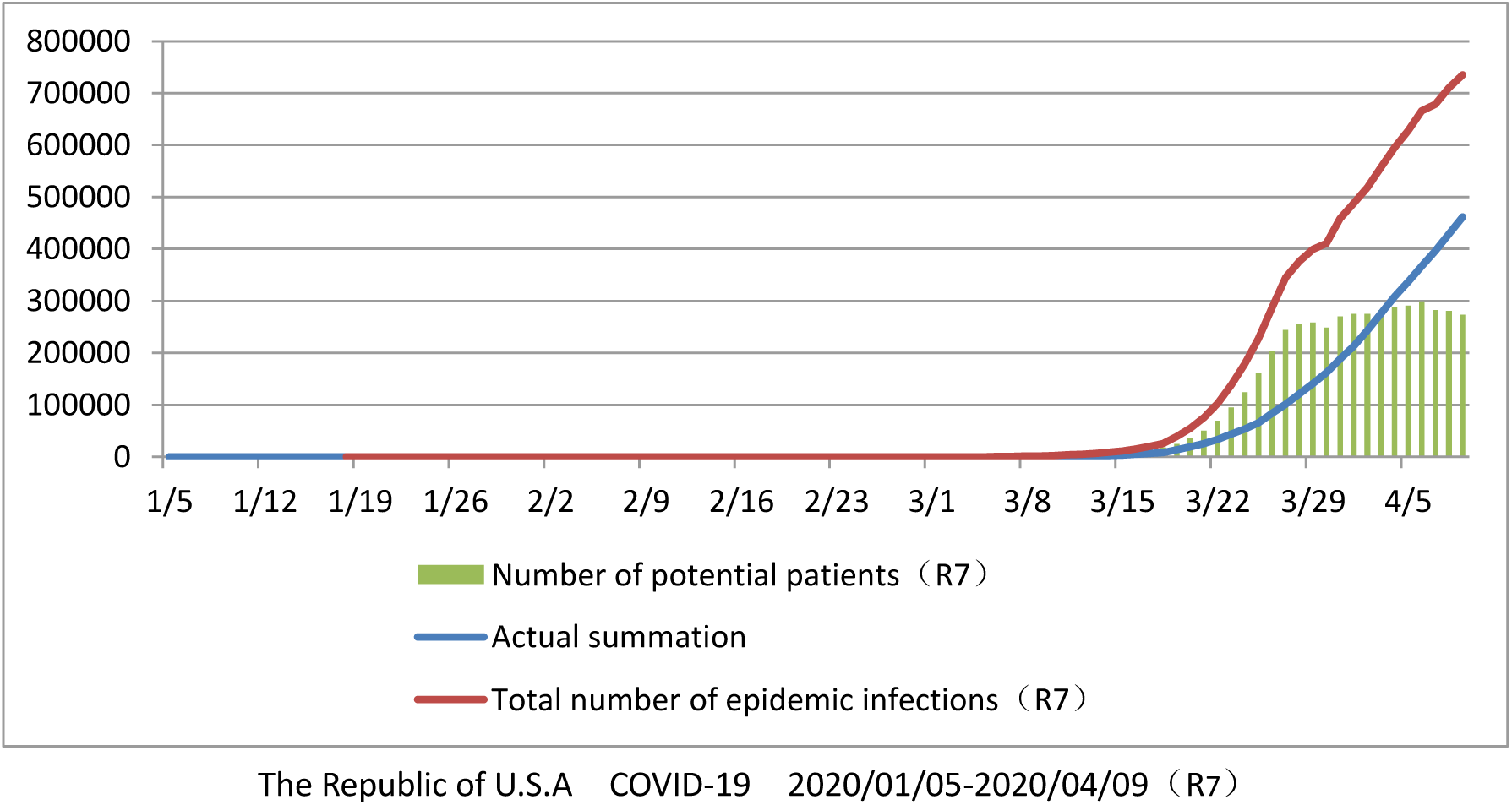

On April 9, 2020, the estimated total number of epidemic infections in the United States reached 735367, far higher than the cumulative total number of 461446 on that day, suggesting that a large number of medical supplies and personnel should be supplemented as soon as possible.

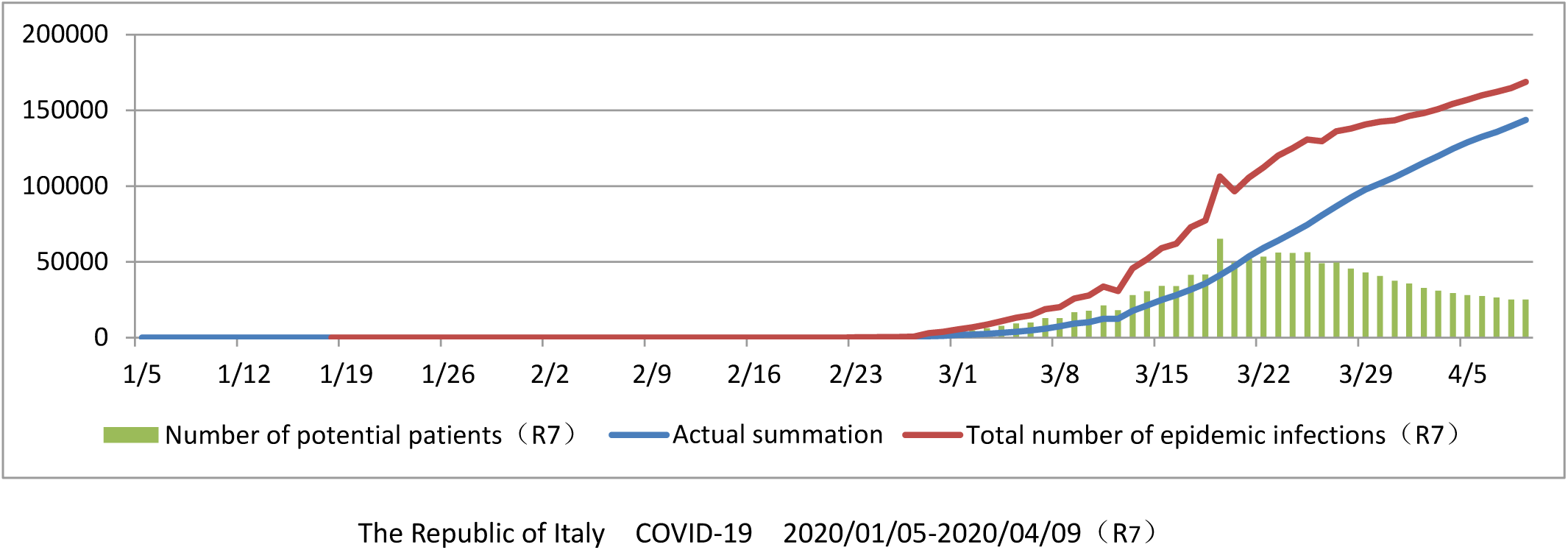

On April 9, 2020, the total number of infectious diseases in Italy was estimated to be 168753, and the slope of the curve slowed down, indicating that the epidemic situation has improved.

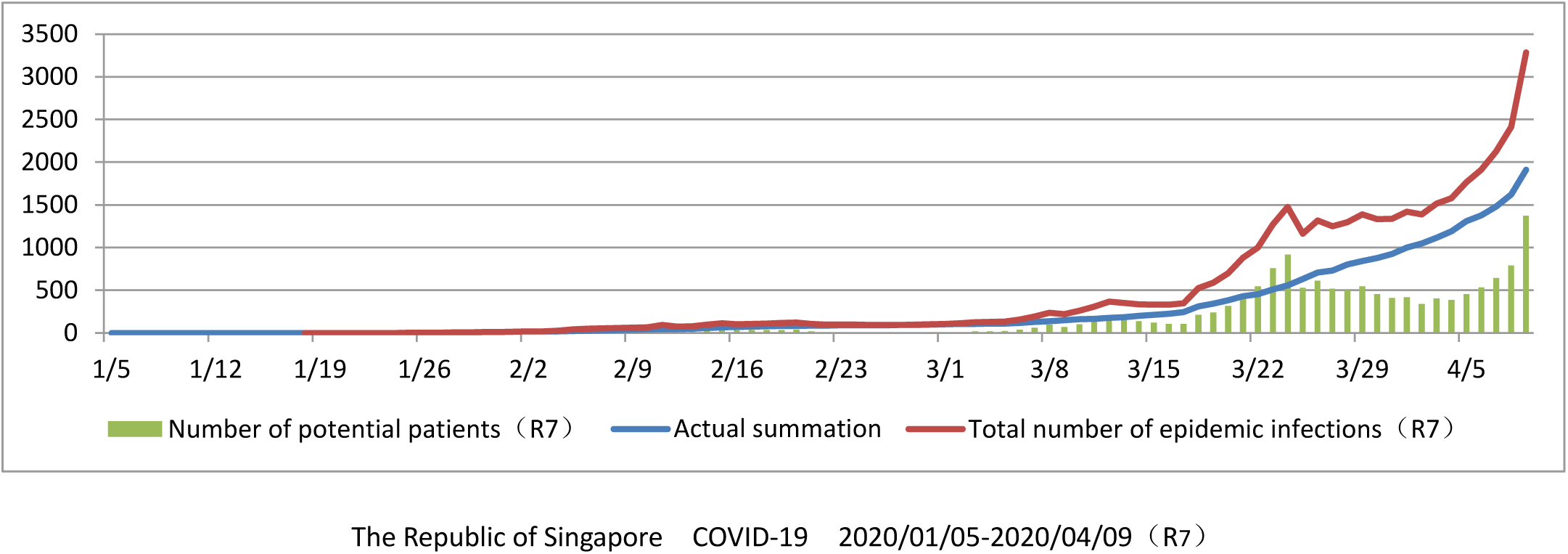

The estimated total number of infections in Singapore fluctuated upward.

On April 7, 2020, more than 100 new cases were reported in Singapore in a single day, marking the outbreak phase. At this time, 74 days have passed since the first patient was found in Singapore (24 January 2020). Good prevention and control measures have provided sufficient time for timely replenishment of medical configuration.

## 3. Summary and extension

### 3.1 Reasonable amendment

Under the unified data statistics standard, the stage of epidemic situation and the potential demand of medical resources in the future can be evaluated by the dynamic curve of M_n_, **R**_n_ and total number of infectious diseases.

In the process of epidemic development, due to various reasons such as detection reagents, personnel, statistics and so on, some stage tests are not sufficient, which can be solved by the calculation of moving average value M_n_.

The national health commission, PRC issued the “novel coronavirus infected pneumonia treatment scheme” trial fifth edition (revised version, February 9, 2020), trial sixth edition (February 19, 2020) statistical standards made two major changes. Among them, on February 12, 2020, 13332 additional clinical cases were confirmed as confirmed cases, which could be concluded by the method described in this paper after relevant data were removed.

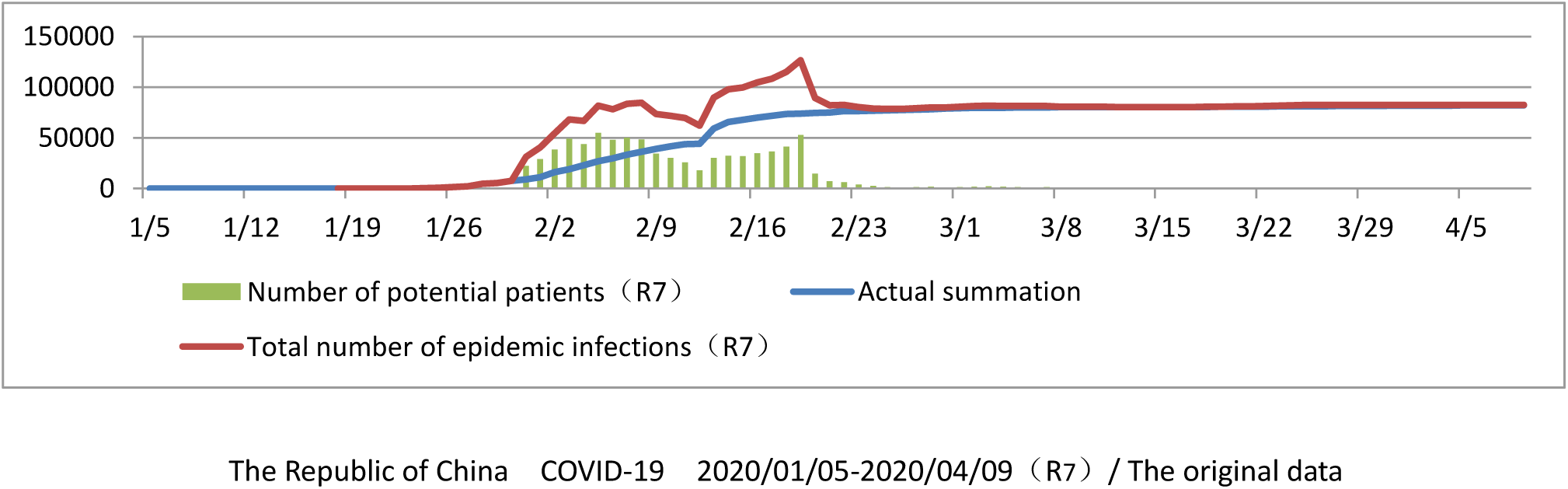

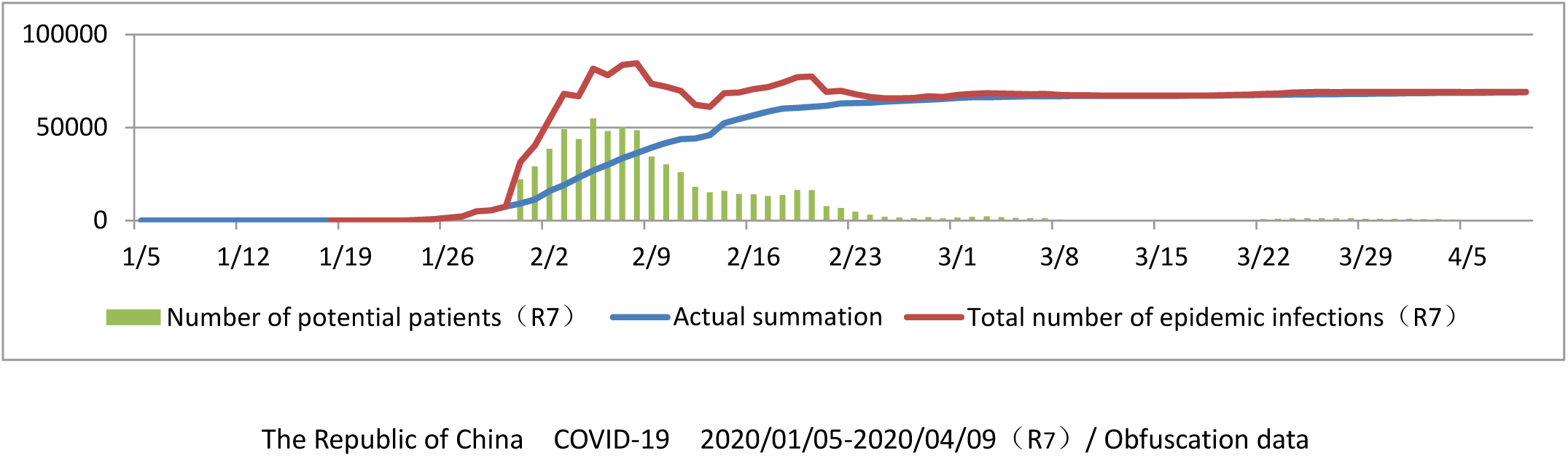

### 3.2 peak prediction

At present, the main prevention and control method of the epidemic in many countries is home quarantine order. Among them, South Korea is the most strict, with the diffusion rate R_7_ dropping rapidly from the upper limit value (R_7_ = 5) for 9 days to below 1, and then maintaining near 1; Italy spent 27 days.

The spread rate R_n_ is a dynamic parameter. The R_7_ fluctuation tracks of Italy and South Korea (starting from R_7_ < 5) are used to predict the epidemic situation in the United States. After 27 days, there will be a difference up to 12 times.

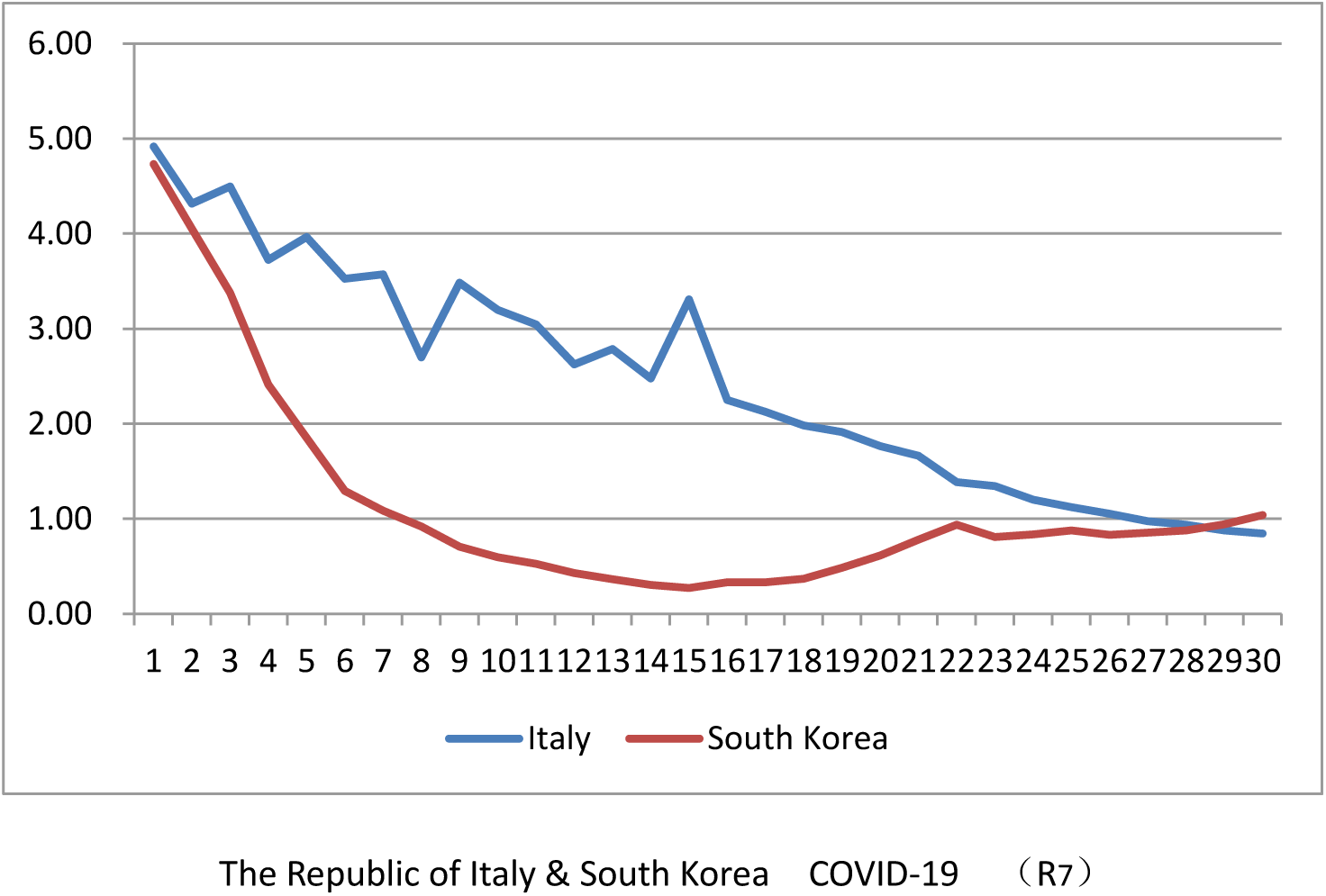

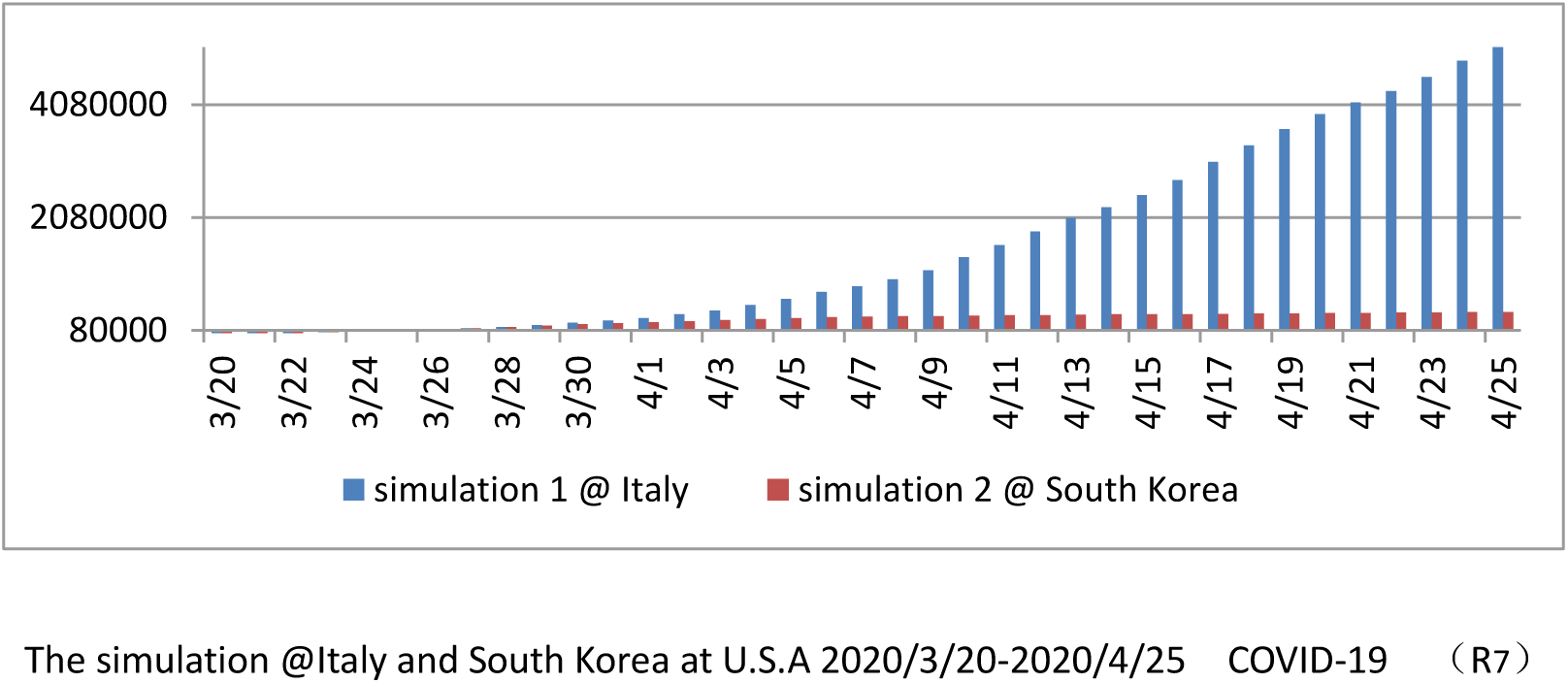

## Data Availability

Reference:
[1] Author: Health Emergency Office of the state health and Health Commission of the people's Republic of China
Title: novel coronavirus pneumonia epidemic situation
Published on the official website of the national health and Health Commission
Year of publication: 2020
[2] By World Health Organization
Title: situation report coronavirus disease 2019 (covid-19)
Published on the official website of the World Health Organization
Year of publication: 2020
[3] Author: Li Baojin; Wu Zilin; Hu Boyong; Chen Hongtao; Liao Silan; Ma CanYe; Liang Huichao;
Title: novel coronavirus infection patients suspected and confirmed by surgical management guidelines
Place of publication: Guangdong medicine (priority)
Published on: February 28, 2020
[4] Data sources: WHO, CDC, ECDC, NHC, DXY, 1point3acres, Worldometers.info, BNO, state and national government health departments, and local media reports, the Johns Hopkins Coronavirus Resource Center .

